# Safety and immunogenicity clinical trial of an inactivated SARS-CoV-2 vaccine, BBV152 (a phase 2, double-blind, randomised controlled trial) and the persistence of immune responses from a phase 1 follow-up report

**DOI:** 10.1101/2020.12.21.20248643

**Authors:** Raches Ella, Siddharth Reddy, Harsh Jogdand, Vamshi Sarangi, Brunda Ganneru, Sai Prasad, Dipankar Das, Dugyala Raju, Usha Praturi, Gajanan Sapkal, Pragya Yadav, Prabhakar Reddy, Savita Verma, Chandramani Singh, Sagar Vivek Redkar, Chandra Sekhar Gillurkar, Jitendra Singh Kushwaha, Satyajit Mohapatra, Amit Bhate, Sanjay Rai, Samiran Panda, Priya Abraham, Nivedita Gupta, Krishna Ella, Balram Bhargava, Krishna Mohan Vadrevu

## Abstract

**Background:** BBV152 is a whole-virion inactivated SARS-CoV-2 vaccine (3 µg or 6 µg) formulated with a Toll-like receptor 7/8 agonist molecule adsorbed to alum (Algel-IMDG). Earlier, we reported findings from a phase 1 (vaccination regimen on days 0 and 14) randomised, double-blind trial on the safety and immunogenicity of three different formulations of BBV152 and one control arm containing Algel (without antigen). Two formulations were selected for the phase 2 (days 0 and 28) study. Here, we report interim findings of a controlled, randomised, double-blind trial on the immunogenicity and safety of BBV152: 3 µg and 6 µg with Algel-IMDG.

**Methods:** We conducted a double-blind, randomised, multicentre, phase 2 clinical trial to evaluate the immunogenicity and safety of BBV152. A total of 380 healthy children and adults were randomised to receive two vaccine formulations (n=190 each) with 3 µg with Algel-IMDG and 6 µg with Algel-IMDG. Two intramuscular doses of vaccines were administered (four weeks apart). Participants, investigators, and laboratory staff were blinded to the treatment allocation. The primary outcome was seroconversion (≥4-fold above baseline) based on wild-type virus neutralisation (PRNT_50_). Secondary outcomes were reactogenicity and safety. Cell-mediated responses were evaluated. A follow-up blood draw was collected from phase 1 participants at day 104 (three months after the second dose).

**Findings:** Among 921 participants screened between Sep 7-13, 2020, 380 participants were randomised to the safety and immunogenicity population. The PRNT_50_ seroconversion rates of neutralising antibodies on day 56 were 92·9% (88·2, 96·2) and 98·3% (95·1, 99·6) in the 3 µg and 6 µg with Algel-IMDG groups, respectively. Higher neutralising titres (2-fold) were observed in the phase 2 study than in the phase 1 study (p<0.05). Both vaccine groups elicited more Th1 cytokines than Th2 cytokines. After two doses, the proportion (95% CI) of solicited local and systemic adverse reactions were 9.7% (6·9, 13·2) and 10.3% (7·4, 13·8) in the 3 µg and 6 µg with Algel-IMDG groups, respectively. No significant difference was observed between the groups. No serious adverse events were reported in this study. Phase 1 follow-up immunological samples at day 104 showed seroconversion in 73·5% (63·6, 81·9), 81·1% (71·4, 88·1), and 73·1% (62·9, 81·8) of individuals in the 3 µg with Algel-IMDG, 6 µg with Algel-IMDG, and 6 µg with Algel groups, respectively.

**Interpretation:** In the phase 1 trial, BBV152 produced high levels of neutralising antibodies that remained elevated in all participants three months after the second vaccination. In the phase 2 trial, BBV152 led to tolerable safety outcomes and enhanced humoral and cell-mediated immune responses. The safety profile of BBV152 is noticeably lower than the rates for other SARS-CoV-2 vaccine platform candidates. The 6 µg Algel-IMDG formulation was selected for the phase 3 efficacy trial.

**Funding:** This work was supported and funded by Bharat Biotech International Limited.

Clinicaltrials.gov: NCT04471519

## Introduction

Severe acute respiratory syndrome coronavirus 2 (SARS-CoV-2), a novel human coronavirus ^1^, has spread worldwide. To date, 194 vaccine candidates are being developed to prevent coronavirus disease 2019 (COVID-19) ^2^. Several such vaccines have been given an Emergency Use Authorization ^3-6^. The virus strain NIV-2020-770 was isolated from a COVID-19 patient, sequenced at the Indian Council of Medical Research-National Institute of Virology (NIV), and provided to Bharat Biotech ^7^. Bio-safety level 3 manufacturing facilities and a well-established Vero cell manufacturing platform aided in the rapid development of BBV152.

Preclinical studies in mice, rats, and rabbits demonstrated appropriate safety profiles and humoral and cell-mediated responses ^8^. Live viral challenge protective efficacy studies in hamsters and nonhuman primates demonstrated rapid viral clearance in the lower and upper respiratory tracts and the absence of lung pathology (after viral challenge) ^9,10^.

Earlier, we reported interim findings from a phase 1 controlled, randomised, double-blind trial on the safety and immunogenicity of three different formulations of BBV152 and one control arm containing Algel (without antigen). This phase 1 trial was successfully conducted with the intention of selecting two formulations for progression to a phase 2 trial. The formulations selected were 3 µg and 6 µg with Algel-IMDG. Here, we report interim findings from a phase 2 controlled, randomised, double-blind trial on the immunogenicity and safety of two formulations of BBV152. Additionally, this paper reports follow-up immunological endpoints from the phase 1 trial (day 104), three months after the second dose.

## Methods

### Trial Design and Participants

This was a randomised, double-blind, multicentre phase 1 trial that was seamlessly followed by a phase 2 trial to evaluate the safety, reactogenicity, tolerability, and immunogenicity of a whole-virion inactivated SARS-CoV-2 vaccine (BBV152) in healthy male and nonpregnant female volunteers across 11 hospitals. Participants were ≥12-<65 years of age at the time of enrolment. At the screening visit, participants were evaluated with both SARS-CoV-2 nucleic acid and serology tests (conducted at a central laboratory using commercially available assays). If individuals were positive for either test, they were excluded from the trial. The median time between the screening visit and vaccination visit was 3 (range: 2-4) days. Participants were screened for eligibility based on their health status, including their medical history, vital signs, and physical examination results and were enrolled after providing signed and dated informed consent forms. Details of the inclusion and exclusion criteria can be found in the protocol.

The trial was conducted across nine sites in nine states in India. The trial was approved by the National Regulatory Authority (India) and the respective Ethics Committees and was conducted in compliance with all International Council for Harmonization (ICH) Good Clinical Practice guidelines. The trial was registered on clinicaltrials.gov: NCT04471519.

### Trial Vaccines

BBV152 (manufactured by Bharat Biotech) is a whole-virion ß-propiolactone-inactivated SARS-CoV-2 vaccine. The vaccine strain NIV-2020-770 contains the D614G mutation, which is characterised by an aspartic acid to glycine shift at amino acid position 614 of the spike protein ^7^.

The candidates were formulated with Algel-IMDG, an imidazoquinoline class molecule (a Toll-like receptor (TLR)7/TLR8 agonist abbreviated as IMDG) adsorbed to Algel. After their eligibility was determined, participants were randomised into two groups: the 3 µg with Algel-IMDG and 6 µg with Algel-IMDG groups. Both vaccines were stored between 2°C and 8°C. All vaccines were stored in a single-use glass vial at a volume of 0·5 mL per dose. The appearance, colour, and viscosity were identical across all formulations.

### Trial Procedures

Vaccines were provided as a sterile liquid that was injected through an intramuscular route (deltoid muscle) at a volume of 0·5 mL/dose in a two-dose regimen on days 0 and 28. No on-site dose preparation was required. Each glass vial contained a single dose of one of the vaccine formulations and required no additional dilution steps. No prophylactic medication (ibuprofen/acetaminophen) was prescribed either before or after vaccination. The follow-up visits were scheduled on days 42, 56, 104, and 194.

In the phase 1 trial, at day 104 (three months after the second dose), 97 (97%), 95 (95%), 92 (92%), and 69 (92%) participants were followed up in the 3 µg with Algel-IMDG, 6 µg with Algel-IMDG, 6 µg with Algel, and Algel alone (control) groups, respectively.

### Randomisation

The master randomisation list was uploaded to the Interactive Web Response System, which contained the randomisation number and intended allocation. The depot manager uploaded the kit code list and assigned the kits to the sites that had the kit codes and the allocation groups. At the site level, the system set the randomisation number and the allotment of the kit without displaying the true group allocation, and the system allocated the same treatment arm for the second visit. A block size of four was utilised. An unblinded Contract Research Organization (CRO), Sclin Soft Technologies, was involved in randomisation for the study.

### Blinding

Participants, investigators, study coordinators, study-related personnel, and the sponsor were blinded to the treatment group allocation (excluding an unblinded CRO that was tasked with the dispatch and labelling of vaccine vials and the generation of the master randomisation code). Participants were assigned a computer-generated randomisation code that maintained blinding. The blinded study nurse was responsible for vaccine preparation and administration. Each vial contained a unique code that ensured appropriate blinding.

### Immunogenicity Assessments

Anti-IgG responses against the spike (S1) protein, receptor-binding domain (RBD), and nucleocapsid (N) protein of SARS-CoV-2 were assessed by enzyme-linked immunosorbent assay (ELISA) and are expressed as geometric mean titres (GMTs). The primary outcome was neutralising antibody titres evaluated by wild-type virus neutralisation assays, namely, (i) a plaque-reduction neutralisation test (PRNT_50_) and (ii) a microneutralisation assay (MNT_50_), at Bharat Biotech. Details of these assays are provided in the Supplementary Appendix.

To compare vaccine-induced responses to natural SARS-CoV-2 infections, 50 convalescent serum samples (collected either one to two months after a nucleic acid test-based diagnosis) were tested by PRNT_50_ and MNT_50_. These serum samples were collected from self-reported symptomatic (n=35) and asymptomatic (n=15) COVID-19 patients and were provided by the NIV, Pune. For symptomatic patients, the ascertainment of severity grading and the requirement for supplemental oxygen was not available. Seroconversion was defined as a postvaccination titre ≥4-fold above the pre-vaccination titre in a participant. All serum samples were analysed in a blinded manner at Bharat Biotech by PRNT_50_ and MNT_50_. To ensure the validity of our assay, a subset of serum samples (n=50) were randomly selected and tested by PRNT_50_ and MNT_50_ at NIV.

Cell-mediated responses were assessed in a subset of participants at three sites on day 42. Serum was used to evaluate Th1 and Th2 dependent antibody isotypes and peripheral blood mononuclear cells (PBMCs) were used to assess the Th1 & Th2 cytokines. The CRO generated a random code containing randomisation numbers, which was provided to the staff to identify participants. Blood (3-5 mL) was collected from participants who consented to have additional blood volume collected on day 42. PBMCs were collected from 58 participants (n=29 each in the 3 µg and 6 µg with Algel-IMDG groups). Pre-vaccination samples collected on day 0 (n=10, from both groups) served as the control. PBMCs collected on day 42 were tested at Indoor Biotechnologies, India, whereas Day 56 PBMCs were tested at Bharat Biotech using Luminex based multiplex assay and Cytokine Bead Array Multiplex Assay (CBA, BD Biosciences, USA), respectively. Luminex based multiplex assay to assessed Th1 (IFN-γ, TNF-α and IL-2) and Th2 (IL-5, IL-10 and IL-13) cytokines. In PBMCs collected on day 104 of the phase 1 trial, T cell memory responses (CD4^+^ CD45RO^+^ T cells and CD4^+^ CD45RO^+^ CD27^+^ T cells) were evaluated at Bharat Biotech. After antigen stimulation of day 104 PBMCs, culture supernatant was collected on day 3, to assess cytokines and secreted SARS-CoV-2 IgG antibodies (by ELISA) on day 6. All samples were analysed in a blinded manner. The details of all assay methods can be found in the Supplementary Appendix.

### Safety Assessments

The secondary outcome was the number and percentage of participants with solicited local and systemic reactogenicity within two hours and seven days after vaccination. Unsolicited adverse events were recorded within 28 days after vaccination.

Participants were observed for two hours postvaccination to assess reactogenicity. They were instructed to record local and systemic reactions within seven days (days 0 to 7 and days 28 to 35) postvaccination using a memory aid. The memory aid contained fields for symptom onset, severity, time to resolution, and concomitant medications and was collected during the next visit to the site. Routine telephone calls were scheduled following the first seven days after each vaccination. Solicited local adverse events included pain at the injection site and swelling, and systemic adverse events included fever, fatigue/malaise, myalgia, body aches, headache, nausea/vomiting, anorexia, chills, generalised rash, and diarrhoea. All unsolicited adverse events were reported by participants throughout the study. Adverse events were graded according to the severity score (mild, moderate, or severe) and whether they were related or unrelated to the investigational vaccine, as detailed in the protocol.

### Sample Size

We assumed that we would observe seroconversion rates (SCRs) of 85% for 3 µg with Algel-IMDG and 95% for 6 µg with Algel-IMDG and a standard deviation (SD) of 0.5 for log_10_ titre. The required sample size for 90% power to find a significant difference (between vaccine formulations differing in the GMT by a ratio of 2) in a trial with a 1:1 allocation using a two-sample z-test at the two-sided 5% significance level was 171 per group. Assuming 10% loss during the study, the number was 190 per group. Sample size estimation was performed using PASS 13 software (Number Cruncher Statistical Systems, USA).

### Statistical Analysis

Safety endpoints are described as frequencies (%). GMTs with 95% confidence intervals (CIs) are presented for immunological endpoints. For continuous variables (below 20 observations), medians and IQRs are reported. The exact binomial calculation was used for the CI estimation of proportions. Wilson’s test was used to test differences in proportions. CI estimation for the GMT was based on the log_10_ (titre) and the assumption that the log_10_ (titre) was normally distributed. A comparison of GMTs was performed with t-tests on the means of the log_10_ (titre). Significance was set at p < 0·05 (2-sided). This preliminary report contains results regarding immunogenicity and safety outcomes (captured on days 0 to 56). Descriptive and inferential statistics were performed using SAS 9·2.

## Results

Among the 921 potential participants screened between Sep 7 and Sep 11, 2020, 380 participants were randomised. Among the 541 initially screened individuals who were excluded, 48 and 123 participants were found to be positive for SARS-CoV-2 with a nucleic acid test and serology, respectively. Due to competitive recruitment, some screened participants (n=188) were eligible but not enrolled and randomised (Figure 1). Other notable exclusions (n=168) were due to inconclusive RT-PCR results. Among enrolled participants, 190 individuals were randomised to each group. The retention rates at day 56 were 96.8% and 93.2% in the 3 µg and 6 µg with Algel-IMDG groups, respectively. Demographic characteristics of participants are presented in Table 1.

**Table 1:**
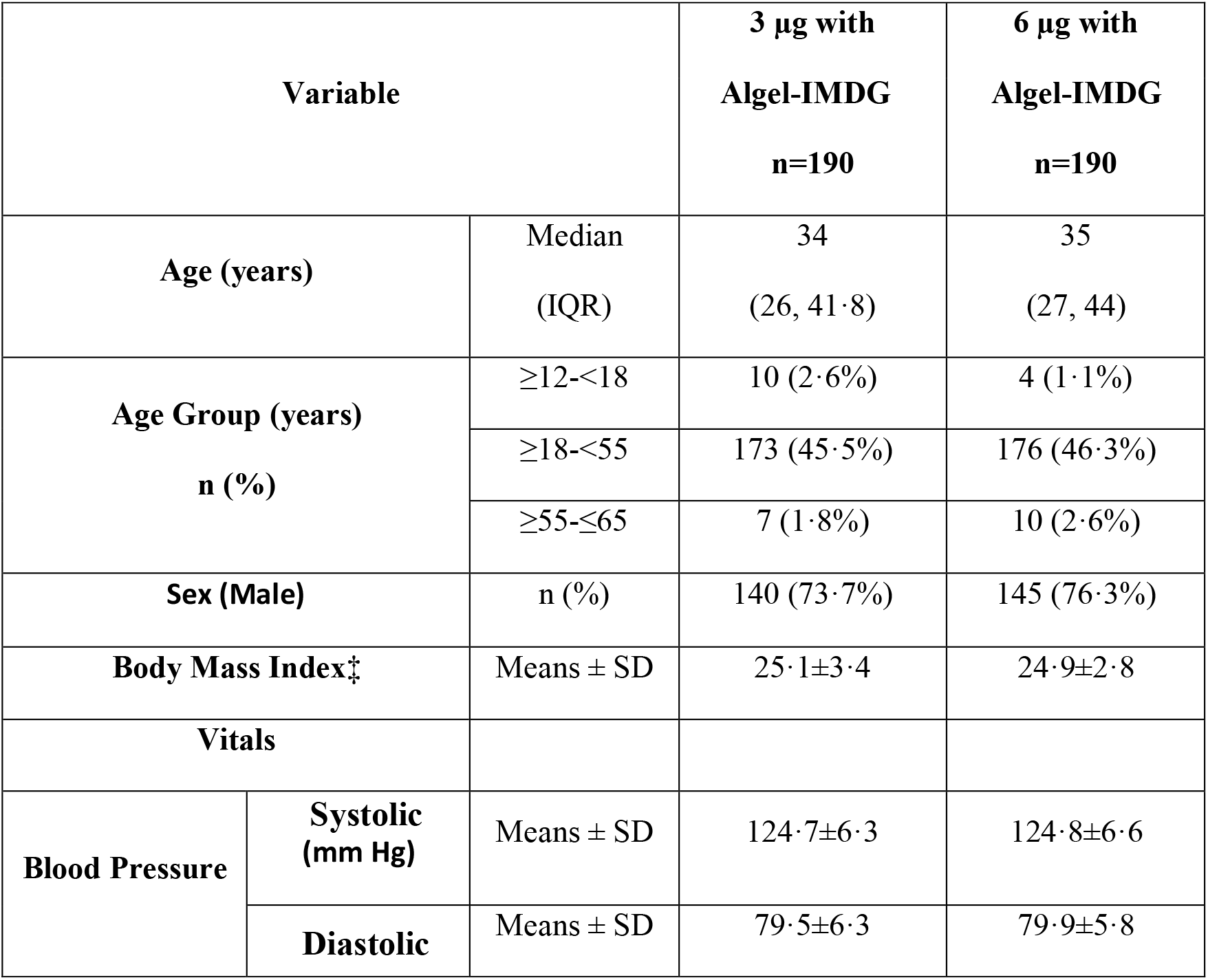
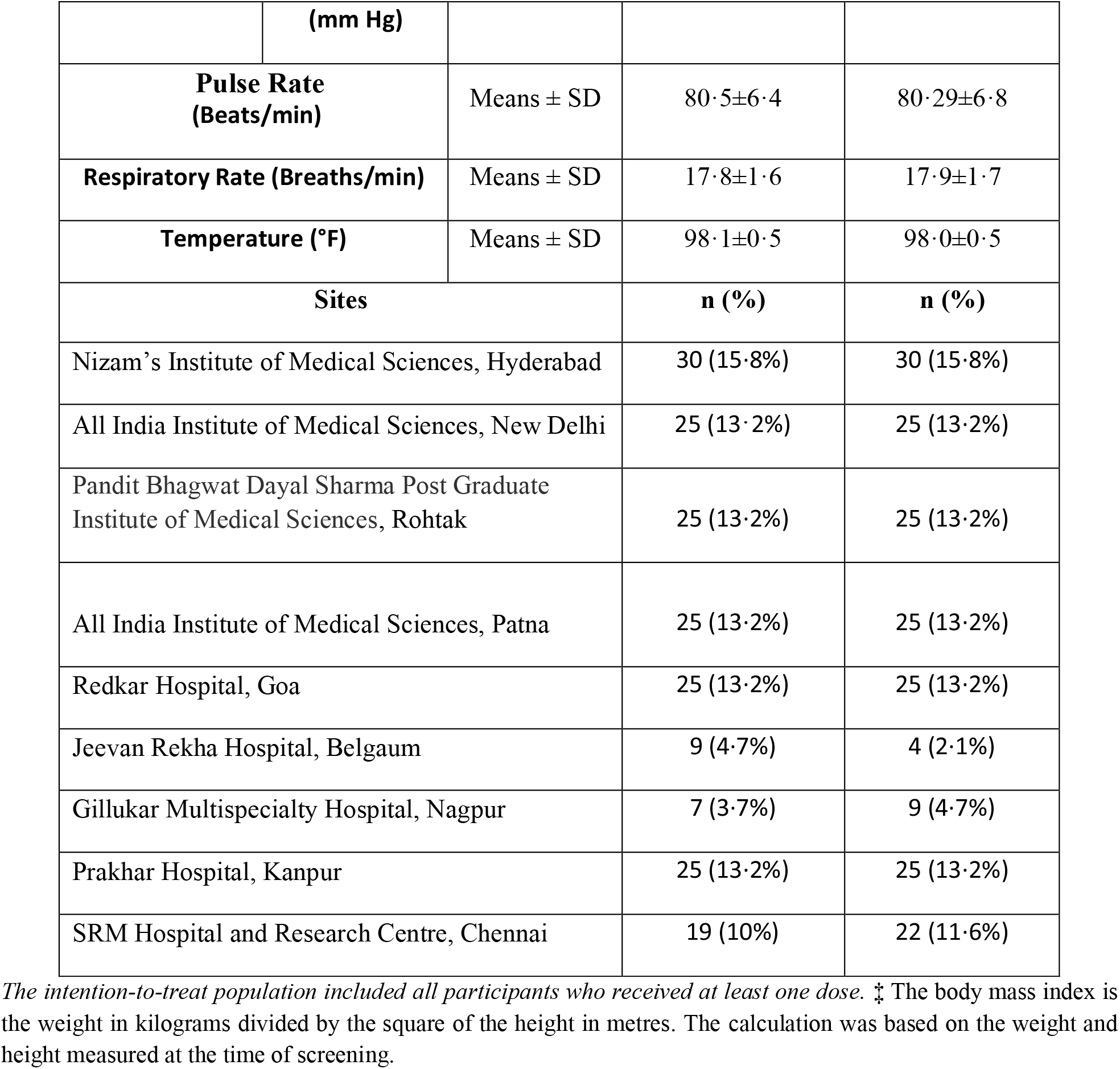
Demographic Characteristics of the Participants in the Intention-to-Treat Population.

**Figure 1:**
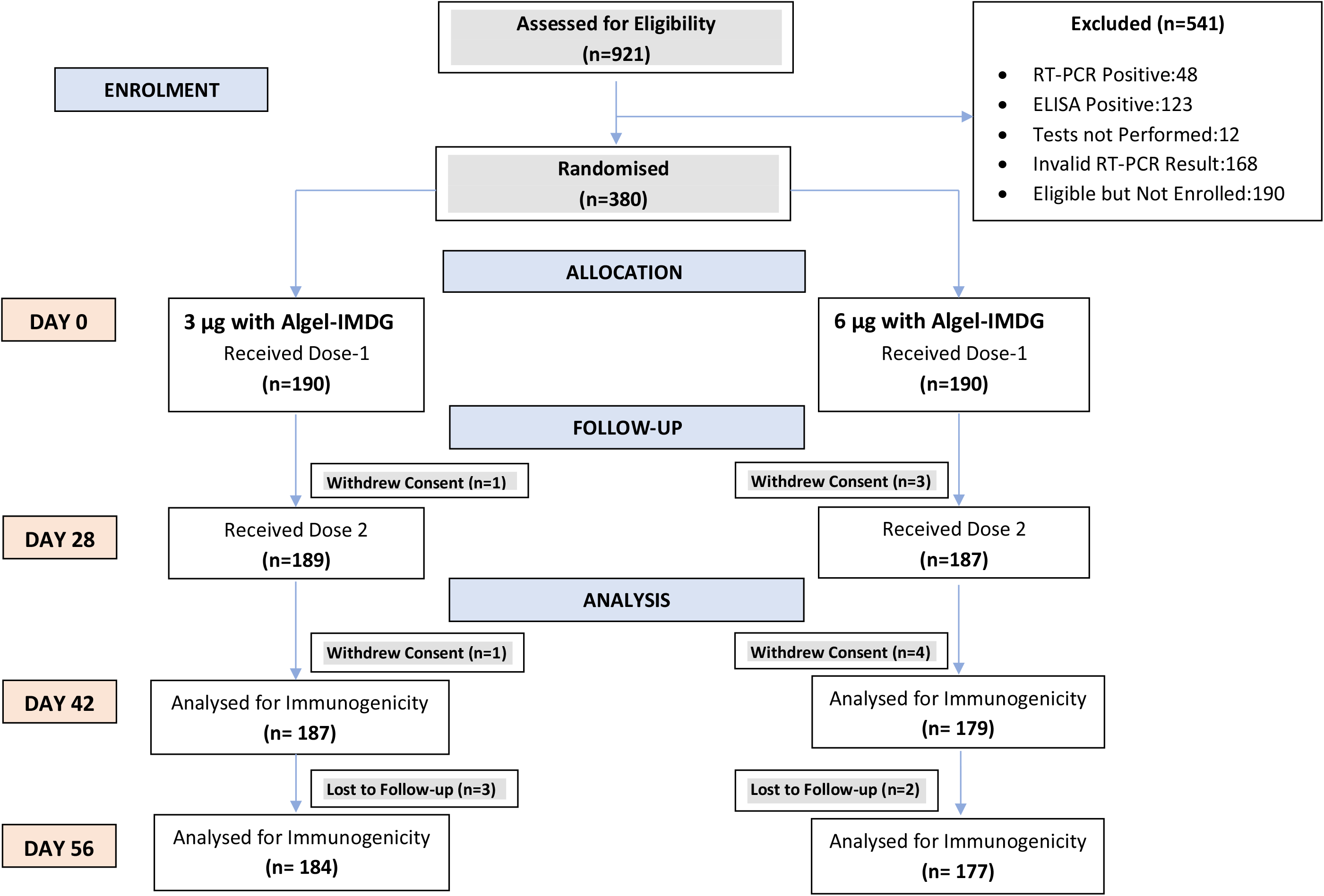
CONSORT.

### Immune Responses

#### Phase 2: Binding Antibody Titres

Binding antibody Anti-IgG titres (GMTs) to all epitopes (S1 protein, RBD, and N protein) increased rapidly after the administration of both doses. Both the 3 µg and 6 µg with Algel-IMDG groups reported comparable anti-S1 protein, -RBD, and -N protein GMTs. The Anti-S1 isotype ratios (IgG1/IgG4) were 2.4 (1.9, 2.9) and 2.1 (1.7, 2.6) in the 3 µg and 6 µg with Algel-IMDG groups, respectively (Table 2).

**Table 2:** SARS-CoV-2 Binding Antibody Responses (Anti-S1, -RBD, and -N IgG)

| **ELISA (Anti-S1, -RBD, and -N IgG)** |  |  | **3 µg with**<br>**Algel-IMDG**<br>**(n=190)** | **6 µg with**<br>**Algel-IMDG**<br>**(n=190)** |
| --- | --- | --- | --- | --- |
| **GMT**<br>**(95% CI)** | **S1-**<br>**Protein** | **Day 0** | 500<br>(500,500) | 500<br>(500,500) |
|  |  | **Day 28** | 2574.2<br>(2228.9, 2973.1) | 2240.5<br>(1942.4, 2584.5) |
|  |  | **Day 42** | 11528.8<br>(10002.7, 13287.8) | 10040.0<br>(8667.0, 11630.5) |
|  |  | **Day 56** | 10413.87<br>(9142.4, 11862.2) | 9541.6<br>(8245.9, 11041.0) |
|  | **RBD-**<br>**Protein** | **Day 0** | 500<br>(500,500) | 500<br>(500,500) |
|  |  | **Day 28** | 1962.7<br>(1726.2, 2231.6) | 2031.6<br>(1777.3, 2322.3) |
|  |  | **Day 42** | 5572.3<br>(4897.5, 6339.9) | 4980.8<br>(4366.7, 5681.3) |
|  |  | **Day 56** | 5874.0<br>(5194.8, 6642.0) | 5558.0<br>(4859.9, 6356.5) |
|  | **N-**<br>**Protein** | **Day 0** | 500<br>(500,500) | 500<br>(500,500) |
|  |  | **Day 28** | 2734.1<br>(2375.1, 3147.5) | 2490.4<br>(2161.7, 2869.2) |
|  |  | **Day 42** | 8957.2<br>(7778.6, 10314.3) | 9211.2<br>(7939.3, 10686.8) |
|  |  | **Day 56** | 8626.0<br>(7528.6, 9883.4) | 8754.0<br>(7589.4, 10097.4) |
| **SCR**<br>**(95% CI)** | **S1-**<br>**Protein** | **Day 28** | 71.20%<br>(64.1, 77.6) | 65.0%<br>(57.5, 72.0) |
|  |  | **Day 42** | 98.4%<br>(95.3, 99.7) | 98.3%<br>(95.1, 99.7) |
|  |  | **Day 56** | 98.4%<br>(95.3, 99.7) | 96.6%<br>(92.8, 98.8) |
|  | **RBD-**<br>**Protein** | **Day 28** | 58.7%<br>(51.2, 65.9) | 58.2%<br>(50.6, 65.6) |
|  |  | **Day 42** | 94.0%<br>(89.6, 97.0) | 93.2%<br>(88.5, 96.5) |
|  |  | **Day 56** | 96.2%<br>(92.3, 98.5) | 94.4%<br>(89.9, 97.3) |
|  | **N-Protein** | **Day 28** | 72.3%<br>(65.2, 78.6) | 71.2%<br>(63.9, 77.7) |
|  |  | **Day 42** | 97.3%<br>(93.8, 99.1) | 95.5%<br>(91.3, 98.0) |
|  |  | **Day 56** | 97.3%<br>(95.3, 100.0) | 96.6%<br>(92.8, 98.8) |
| **Isotype Mean (95% CI)** | **Day 28** |  | 1.7<br>(1.3, 2.1) | 1.9<br>(1.5, 2.8) |
|  | **Day 42** |  | 2.4<br>(1.9, 2.9) | 2.2<br>(1.7, 2.6) |

Binding antibody results at baseline (day 0), 4 weeks after the first vaccination (day 28), 2 weeks after the second vaccination (day 42), and 4 weeks after the second vaccination (day 56) for the 3 µg (n=190) and 6 µg (n=190) with Algel-IMDG groups are shown. IgG titres against anti-S1, anti-RBD, anti-N, and the anti-S1 IgG1/IgG4 ratio. The cut-off for detectable antibodies was set at 1:500. Endpoint titre dilution for day 28 serum samples was determined with baseline (day 0) and interpolated from the raw optical density (OD) data of the corresponding day 0 sample. The cut-off (mean±3 SD) for day 0 was calculated based on the absorbance of all serum dilutions (1:500 to 32000) tested, except the lowest dilution (1:500). Isotyping titres on day 56 were not analysed. SCRs were defined based on the proportion of titres that increased ≥4-fold compared to baseline.

#### Phase 2: Neutralising Antibody Titres (at day 56, four weeks after the second dose)

GMTs (PRNT_50_) were 100·9 (74·1, 137·4) and 197·0 (155·6, 249·4) in the 3 µg and 6 µg with Algel-IMDG groups, respectively. The GMT in the 6 µg with Algel-IMDG group was higher and found to be significantly different than that in the 3 µg with Algel-IMDG group. The 6 µg with Algel-IMDG-induced responses were comparable to those observed in convalescent serum collected from patients who had recovered from COVID-19 (Figure 2A). The proportions of participants who experienced seroconversion based on PRNT_50_ (95% CI) were 92·9% (88·2, 96·2) and 98·3% (95·1, 99·6) in the 3 µg and 6 µg with Algel-IMDG groups, respectively (Figure 2B). GMTs (MNT_50_) were 92·5 (77·7, 110·2) and 160·1 (135·8, 188·8) in the 3 µg and 6 µg with Algel-IMDG groups, respectively (Figure 2C). The proportions of participants who experienced seroconversion based on MNT_50_ (95% CI) were 88·0% (82·4, 92·3) and 96·6% (92·8, 98·8) in the 3 µg and 6 µg with Algel-IMDG groups, respectively (Figure 2D and Table S2 in the Supplementary Appendix). The PRNT_50_ and MNT_50_ GMTs in the 6 µg with Algel-IMDG group were higher and significantly different than those in the 3 µg with Algel-IMDG group.

**Figure 2:**
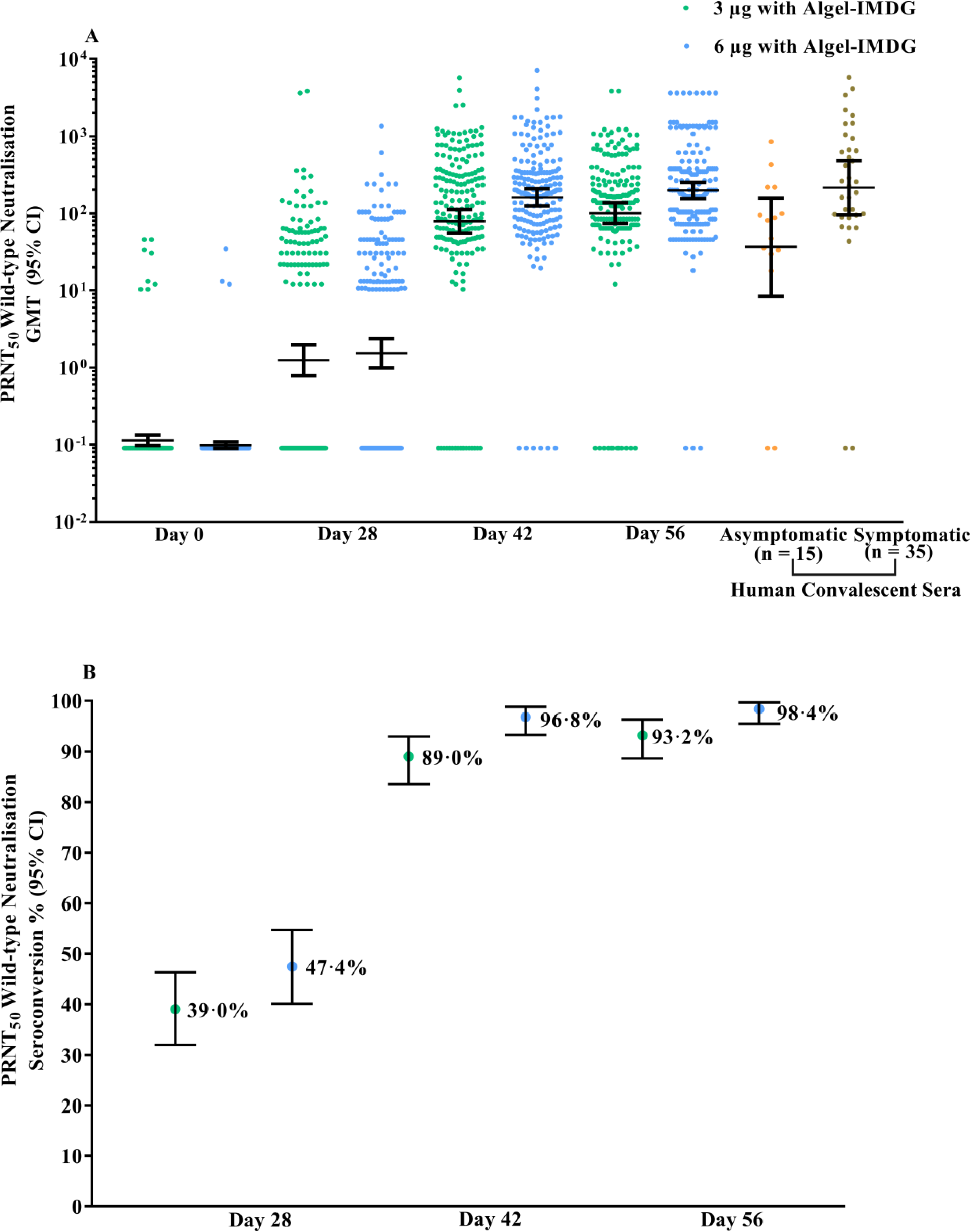
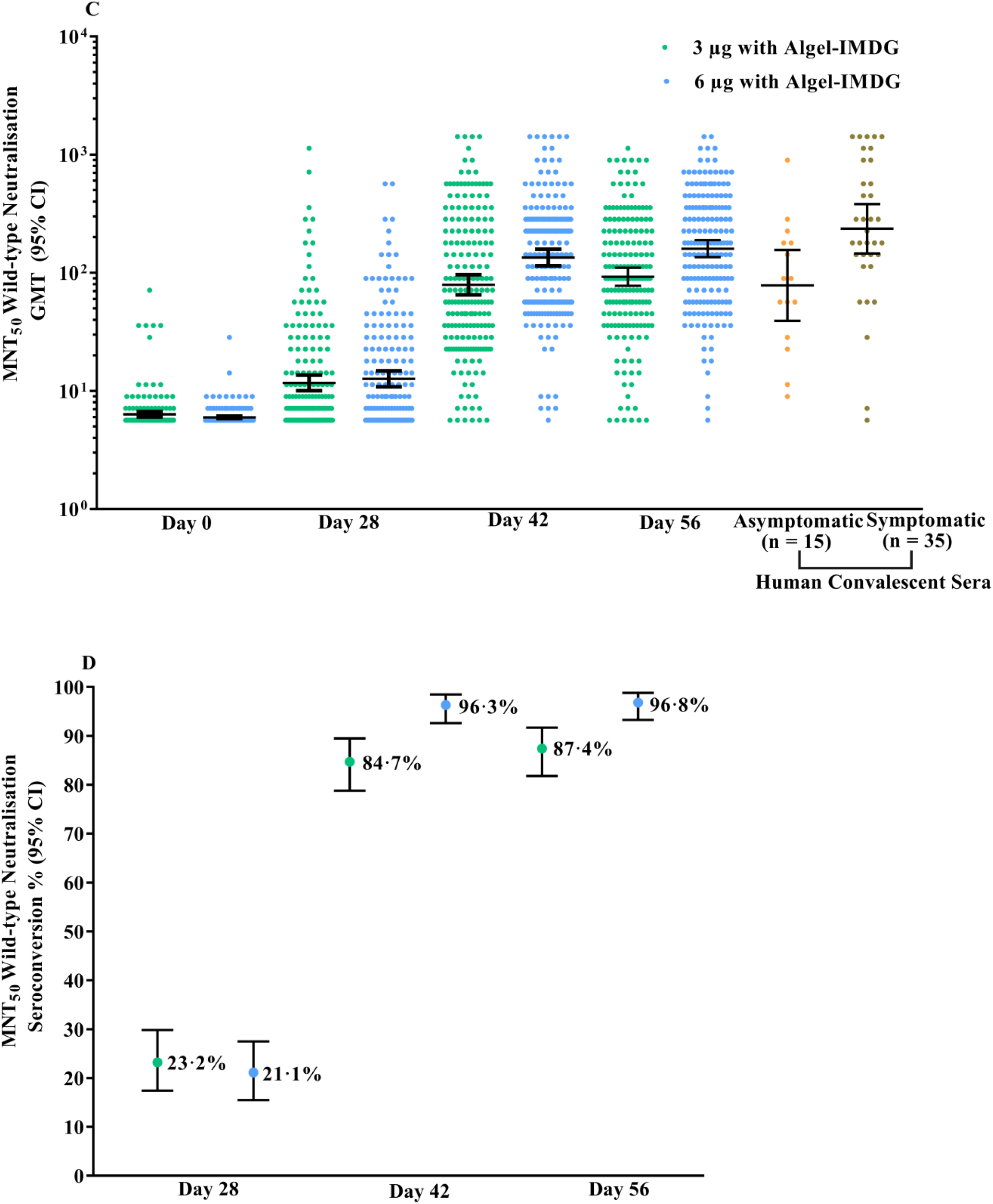
SARS-CoV-2 Neutralising Antibody Responses. Titres of the wild-type SARS-CoV-2 neutralisation assay (PRNT_50_ and MNT_50_) at baseline (day 0), 4 weeks after the first vaccination (day 28), 2 weeks after the second vaccination (day 42), and 4 weeks after the second vaccination (day 56) for the 3 µg (n=190) and 6 µg (n=190) with Algel-IMDG groups are shown. SCRs were defined based on the proportion of titres ≥4-fold above baseline. The dots and horizontal bars represent the SCR and 95% CI, respectively (panels A&C). In panels B&D, the dots and horizontal bars represent individual data points and the geometric mean (95% CI). The human convalescent serum (HCS) panel included specimens from PCR-confirmed symptomatic/asymptomatic COVID-19 participants obtained at least 30-60 days after diagnosis (n=50 samples).

PRNT_50_ wild-type neutralising antibody responses for a subset of paired serum samples (n=50) were analysed at NIV and Bharat Biotech (on day 42, 2 weeks after the second vaccination in both groups). In comparisons of PRNT_50_ assays between laboratories, a strong agreement was observed. Seroconversion in any three age groups was always found to be above 90%. No significant differences were observed in seroconversion and GMTs across the three age groups and between both sexes, but small numbers of samples were included in the ≥12-<18 and ≥55-<65 age groups (Table S3 in the Supplementary Appendix).

#### Phase 1: Neutralising Antibody Titres (at day 104, three months after the second dose)

GMTs (MNT_50_) were 39·9 (32·0, 49·9), 69·5 (53·7, 89·9), and 53·3 (40·1, 71·0) in the 3 µg with Algel-IMDG, 6 µg with Algel-IMDG and 6 µg with Algel groups, respectively (Figure 3A). The proportions of participants who experienced seroconversion based on MNT_50_ (95% CI) were 73·5% (63·6, 81·9), 81·1% (71·4, 88·1), 73·1% (62·9, 81·8) in the 3 µg with Algel-IMDG, 6 µg with Algel-IMDG, and 6 µg with Algel groups, respectively (Figure 3B). SCRs and GMT responses in the 6 µg with Algel-IMDG group were higher and were significantly different than those in the 3 µg with Algel-IMDG and 6 µg with Algel groups (Table S4 in the Supplementary Appendix). In the 6 µg with Algel-IMDG group, there were no significant differences in SCRs and GMTs between day 42 (two weeks after the second dose) and 104 (three months after the second dose). The phase 2 neutralisation GMTs were higher and significantly different than those in phase 1 (Figure 3C). At four weeks after the second dose of 6 µg with Algel-IMDG, the MNT _50_ GMT ratio between Phase 1 and 2 was 1.9 (95%CI: 1·5, 2·6).

**Figure 3:**
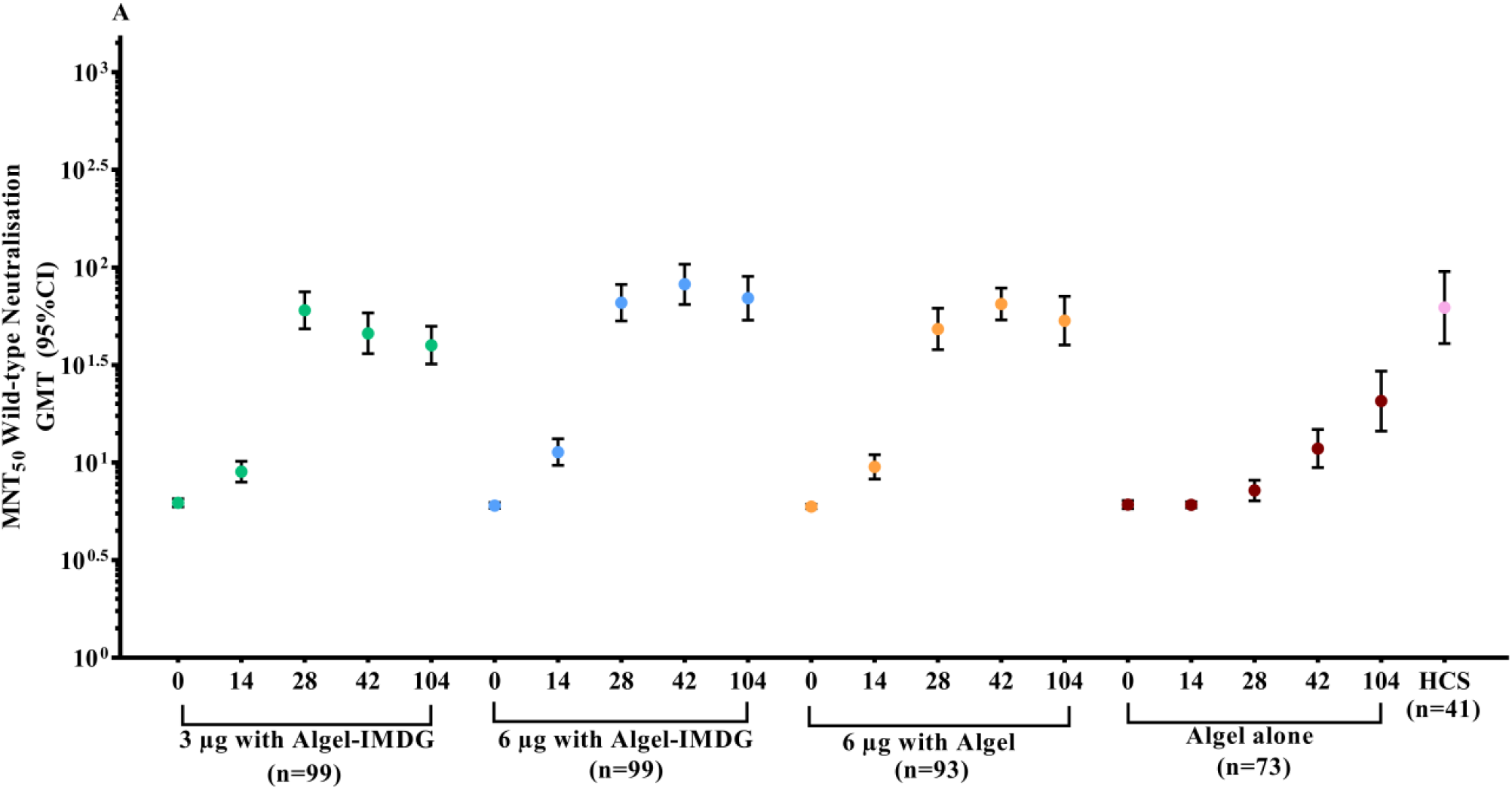
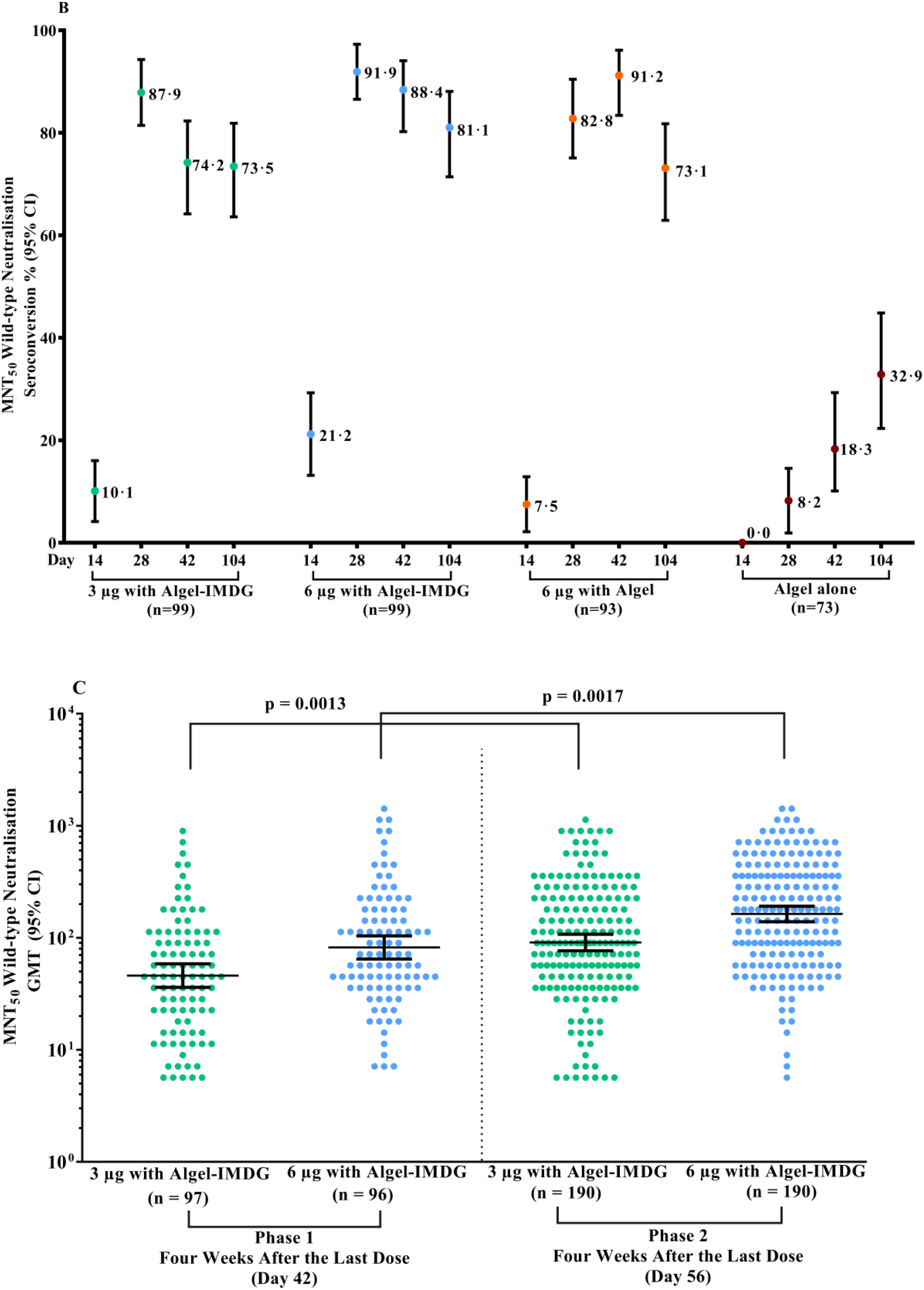
Neutralising Responses from Phase 1 and 2 Trials. Panels A & B show phase 1 GMTs of the wild-type SARS-CoV-2 MNT_50_ at baseline (day 0), 2 weeks after the second vaccination (day 28), 4 weeks after the second vaccination (day 42), and 3 months after the second vaccination (day 104) for the 3 µg and 6 µg with Algel-IMDG groups, the 6 µg with Algel group, and the Algel-only control arm. In the phase 1 trial, the dosing schedule was days 0 and 14 for the first and second doses of the vaccine, respectively. SCRs were defined based on the proportion of titres ≥4-fold above baseline. The HCS panel included specimens from PCR-confirmed symptomatic/asymptomatic COVID-19 participants obtained at least 30 days after diagnosis (41 samples for MNT_50_). In the phase 2 trial, the dosing schedule was days 0 and 28 for the first and second doses of the vaccine, respectively. Panel C shows phase 1 and 2 GMT) of the wild-type SARS-CoV-2 MNT_50_. GMTs in phase 2 were significantly higher than those in phase 1.

#### Cell-mediated Responses

##### Phase 2 (at day 42, two weeks after the second dose)

The ratios of Th1/Th2 cytokines (IFN-γ + TNF-α + IL-2 /IL-5 + IL-13 + IL-10) were biased to a Th1 response (Figure 4A). Th2 responses were detected at minimal levels in both formulations, as observed by IL-5, IL-10 and IL-13 responses (Figure 4B).

**Figure 4:**
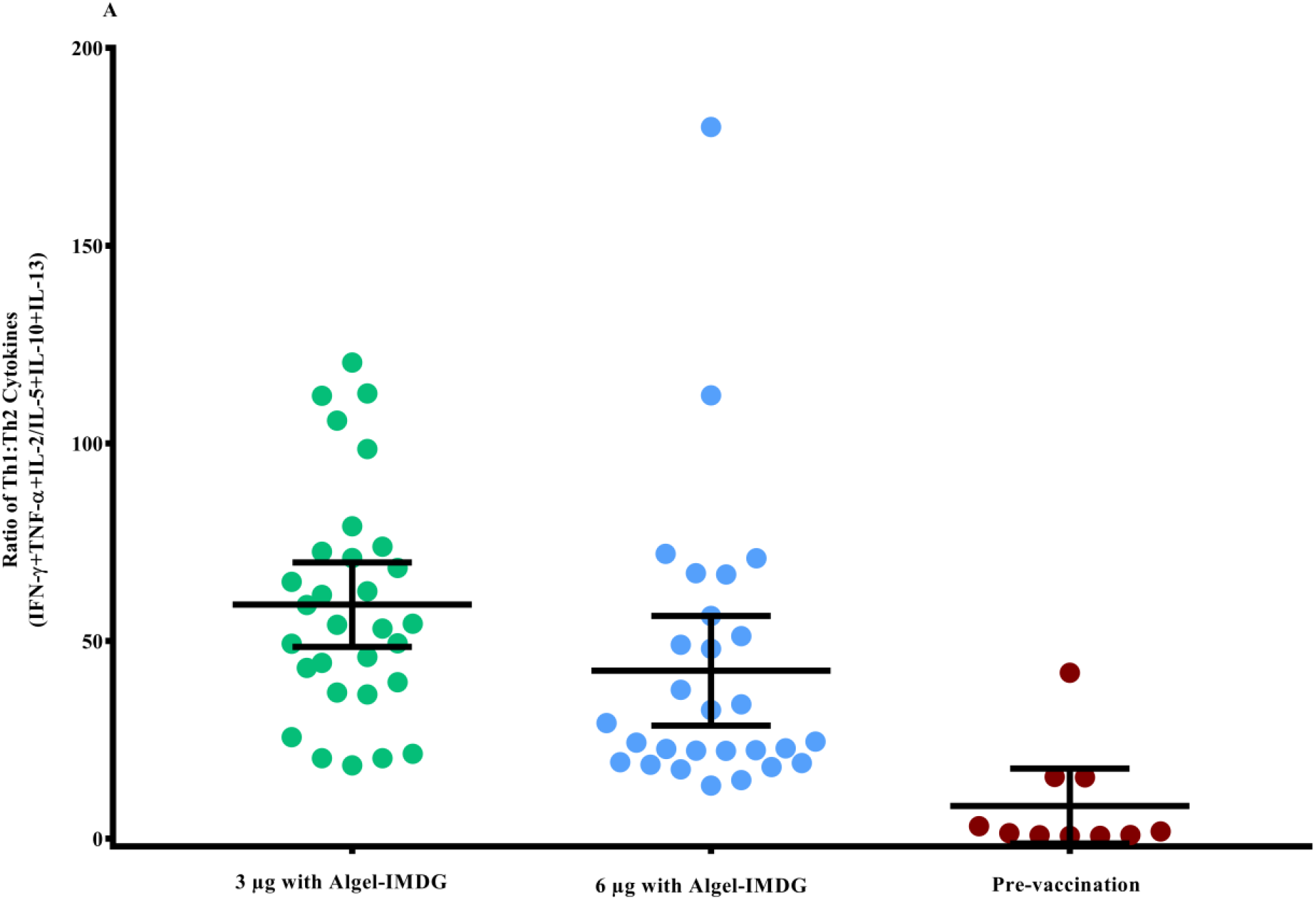
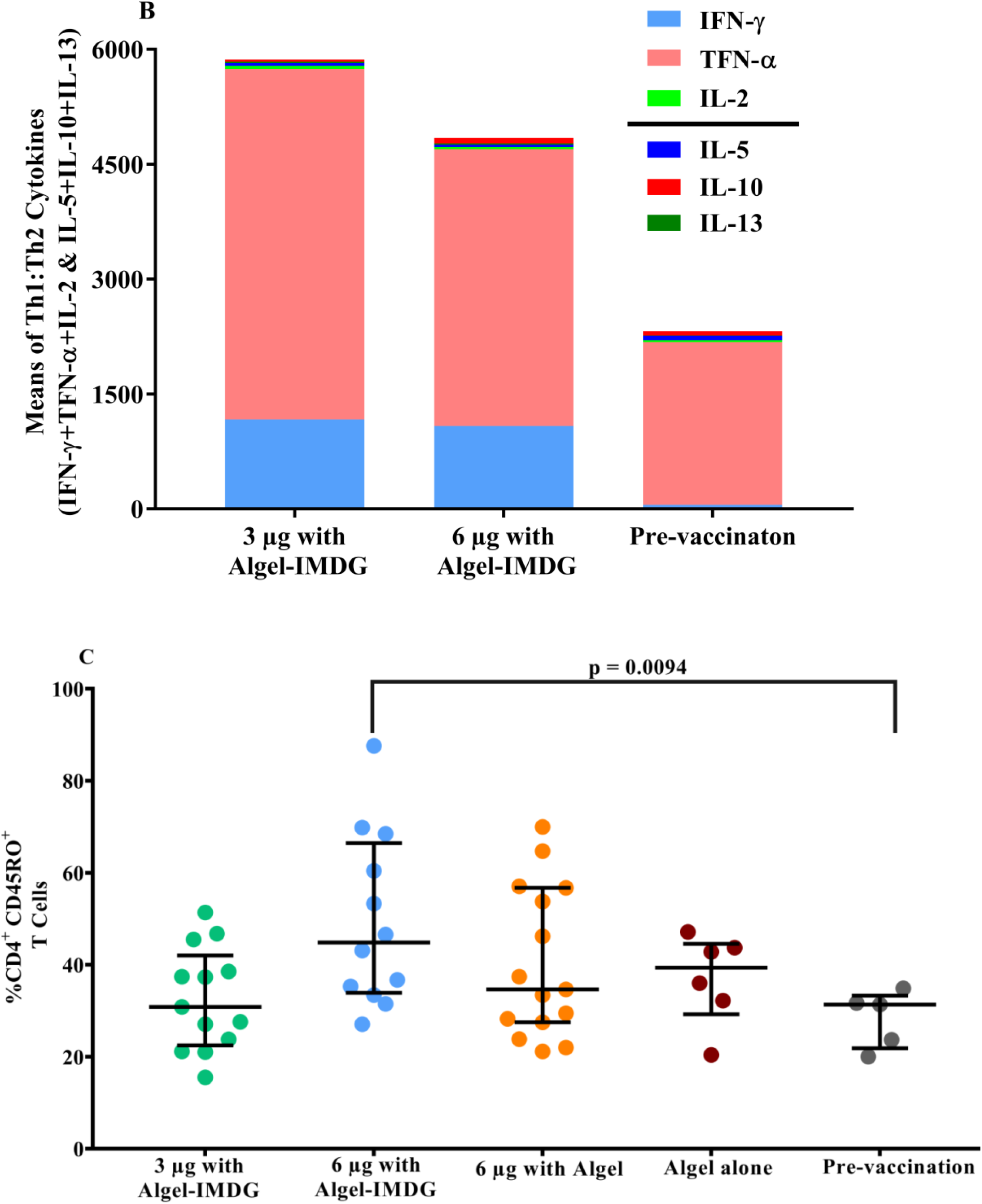
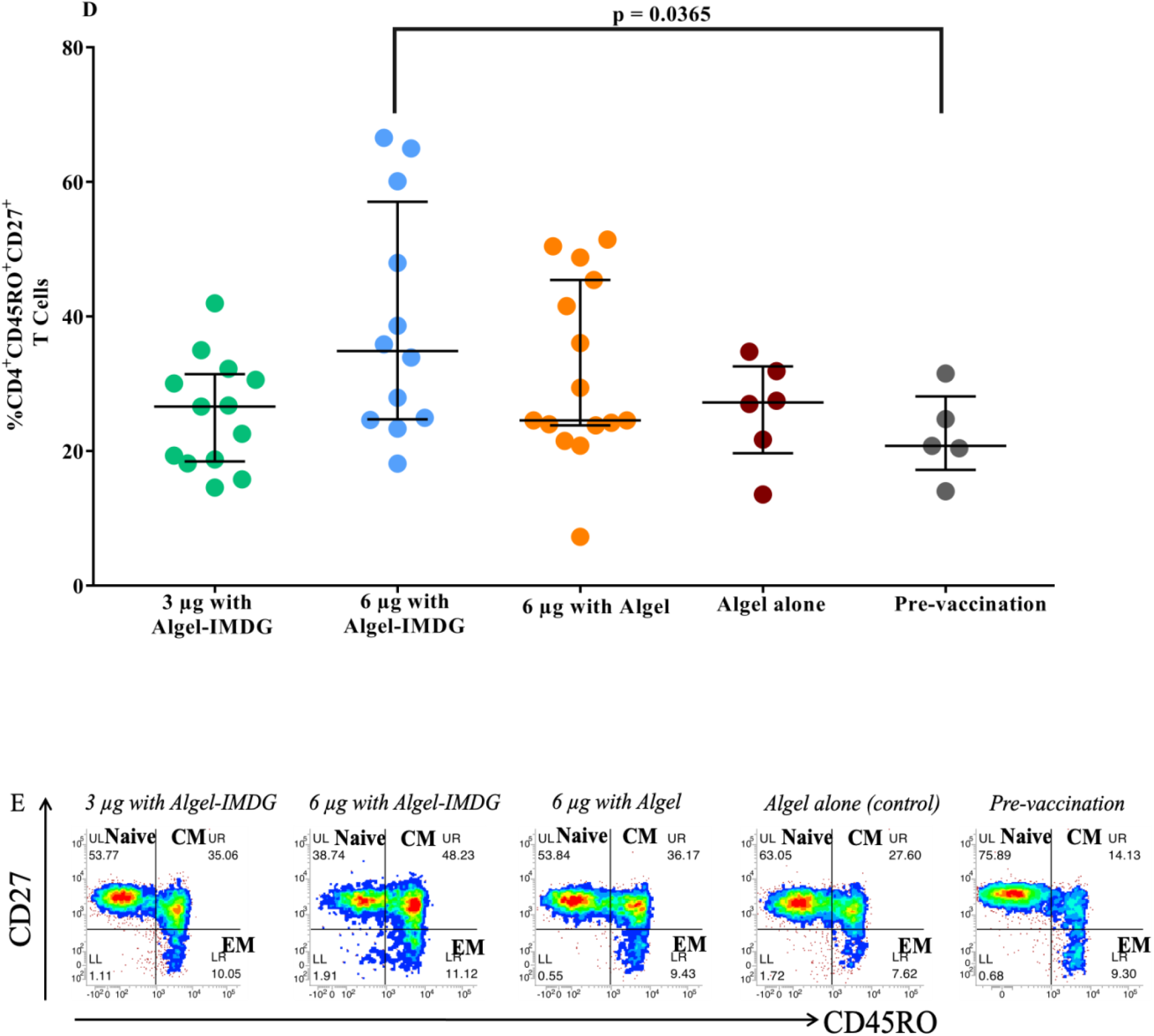
SARS-CoV-2 Cell-mediated Responses. Cytokine levels in day supernatants from 58 participants (n=29 in each of the 3 µg and 6 µg with Algel-IMDG groups) and controls (n=10 pre-vaccination samples from both groups) with proliferative responses to BBV152 vaccination whose PBMCs were evaluated after stimulation with SARS-CoV-2 peptides are shown. Samples were collected two weeks after the second vaccination (day 42) for the 3 µg and 6 µg with Algel-IMDG groups. Error bars show the mean (95% CI) of the ratio of Th1/Th2 cytokines: [interferon-gamma (IFN-γ) + IL-2]/[IL-5+IL-13] (panel A). Th1 and Th2 cytokines are represented by stacked bars (panel B). Panels C & D: Scatter plot represents the frequencies of antigen-specific T cell memory responses 3 months after the second vaccination (day 104) for the 3 µg and 6 µg with Algel-IMDG groups, 6 µg with Algel group, and the Algel-only control arm from the phase 1 trial participants are shown. Dots represent an individual data point with medians and IQR. Panel E: Representative dot plots from one participant, representative of the group mean value. Gating was done on CD4^+^ T cells illustrating the frequencies of naïve effector memory (_EM_) T_EM_, CD45RO-CD27^+^, central memory (_CM_) T_CM_, CD45RO^+^CD27-, and T_EM_, CD45RO^+^CD27^+^ CD4^+^ T cells.

##### Phase 2 (at day 56, two weeks after the second dose)

We observed a profound increase in the levels of Th1-biased cytokines, such as IFN-γ, IL-2 and TNF-α responses on day 56, performed by the CBA method (Supplementary Figure S1).

##### Phase 1 (at day 104, three months after the second dose)

In the phase 1 trial, PBMCs from a subset of participants at one site were collected to evaluate T cell memory responses at day 104. Formulations with Algel-IMDG generated a T cell memory response, as shown by an increase in the frequency of effector memory CD4^+^ CD45RO^+^ T cells and CD4^+^ CD45RO^+^ CD27^+^ T cells compared to pre-vaccination samples (Figure 4C & D). Placebo samples also showed a T cell memory response. We also detected secreted IgG antibodies in the cell culture supernatant by ELISA, and the antibody titre ranged from neat (undiluted) to 1:64 (Supplementary Table S5). Further effector function of activated and differentiated T cells was demonstrated by the measurement of Th1 mediated cytokines (Supplement Table S6).

### Reactogenicity

After dose 1, the proportions of solicited local adverse reactions (95% CI) reported were 4·7% (2·2, 8·8) and 4·2% (1·8, 8·1) in the 3 µg and 6 µg with Algel-IMDG groups, respectively. The proportions of solicited systemic adverse reactions (95% CI) were 4·7% (2·2, 8·8) and 7·4% (4·1, 12·1) in the 3 µg and 6 µg with Algel-IMDG groups, respectively (Table 3). After both doses, the most common solicited adverse events were injection site pain, at 2·6% (0·9, 6·0) and 3·2 (1·2, 6·8) in the 3 µg and 6 µg with Algel-IMDG groups, respectively. The majority of the adverse events were mild and resolved within 24 hours of onset. After both doses, the proportions (95% CI) of solicited local and systemic adverse reactions were 9·7% (6·9, 13·2) and 10·3% (7·4, 13·8) in the 3 µg and 6 µg with Algel-IMDG groups, respectively. No significant differences were observed between the groups.

**Table 3:**
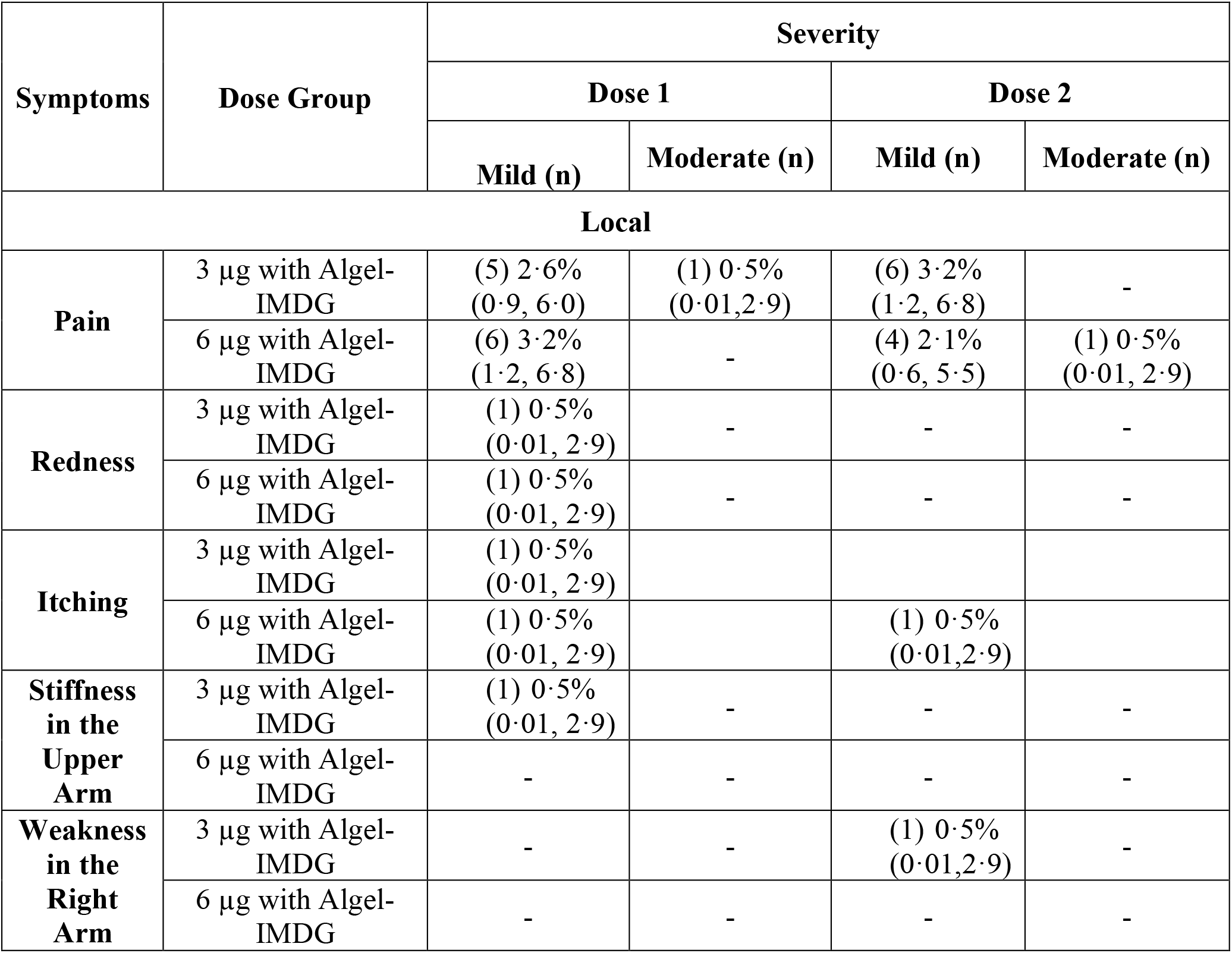
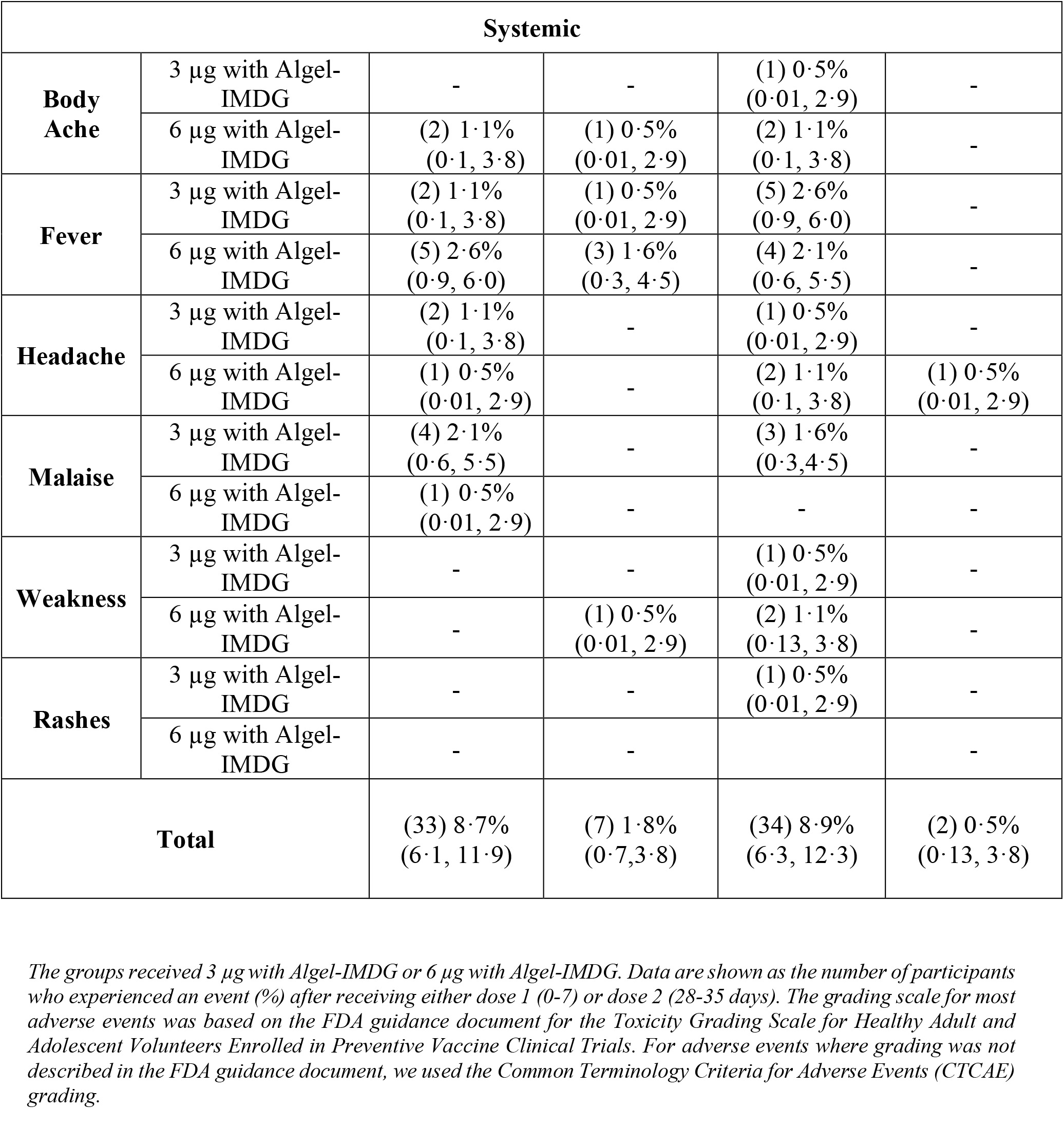
Solicited Adverse Events After Two Doses in the Safety Set.

| Symptoms | Dose Group | Severity |  |  |  |
| --- | --- | --- | --- | --- | --- |
|  |  | Dose 1 |  | Dose 2 |  |
|  |  | Mild (n) | Moderate (n) | Mild (n) | Moderate (n) |
| Local |  |  |  |  |  |
| Pain | 3 µg with Algel-IMDG | (5) 2·6%<br>(0·9, 6·0) | (1) 0·5%<br>(0·01,2·9) | (6) 3·2%<br>(1·2, 6·8) | - |
|  | 6 µg with Algel-IMDG | (6) 3·2%<br>(1·2, 6·8) | - | (4) 2·1%<br>(0·6, 5·5) | (1) 0·5%<br>(0·01, 2·9) |
| Redness | 3 µg with Algel-IMDG | (1) 0·5%<br>(0·01, 2·9) | - | - | - |
|  | 6 µg with Algel-IMDG | (1) 0·5%<br>(0·01, 2·9) | - | - | - |
| Itching | 3 µg with Algel-IMDG | (1) 0·5%<br>(0·01, 2·9) |  |  |  |
|  | 6 µg with Algel-IMDG | (1) 0·5%<br>(0·01, 2·9) |  | (1) 0·5%<br>(0·01,2·9) |  |
| Stiffness in the Upper Arm | 3 µg with Algel-IMDG | (1) 0·5%<br>(0·01, 2·9) | - | - | - |
|  | 6 µg with Algel-IMDG | - | - | - | - |
| Weakness in the Right Arm | 3 µg with Algel-IMDG | - | - | (1) 0·5%<br>(0·01,2·9) | - |
|  | 6 µg with Algel-IMDG | - | - | - | - |

| Systemic |  |  |  |  |  |
| --- | --- | --- | --- | --- | --- |
| **Body Ache** | 3 µg with Algel-IMDG | - | - | (1) 0.5%<br>(0.01, 2.9) | - |
|  | 6 µg with Algel-IMDG | (2) 1.1%<br>(0.1, 3.8) | (1) 0.5%<br>(0.01, 2.9) | (2) 1.1%<br>(0.1, 3.8) | - |
| **Fever** | 3 µg with Algel-IMDG | (2) 1.1%<br>(0.1, 3.8) | (1) 0.5%<br>(0.01, 2.9) | (5) 2.6%<br>(0.9, 6.0) | - |
|  | 6 µg with Algel-IMDG | (5) 2.6%<br>(0.9, 6.0) | (3) 1.6%<br>(0.3, 4.5) | (4) 2.1%<br>(0.6, 5.5) | - |
| **Headache** | 3 µg with Algel-IMDG | (2) 1.1%<br>(0.1, 3.8) | - | (1) 0.5%<br>(0.01, 2.9) | - |
|  | 6 µg with Algel-IMDG | (1) 0.5%<br>(0.01, 2.9) | - | (2) 1.1%<br>(0.1, 3.8) | (1) 0.5%<br>(0.01, 2.9) |
| **Malaise** | 3 µg with Algel-IMDG | (4) 2.1%<br>(0.6, 5.5) | - | (3) 1.6%<br>(0.3, 4.5) | - |
|  | 6 µg with Algel-IMDG | (1) 0.5%<br>(0.01, 2.9) | - | - | - |
| **Weakness** | 3 µg with Algel-IMDG | - | - | (1) 0.5%<br>(0.01, 2.9) | - |
|  | 6 µg with Algel-IMDG | - | (1) 0.5%<br>(0.01, 2.9) | (2) 1.1%<br>(0.13, 3.8) | - |
| **Rashes** | 3 µg with Algel-IMDG | - | - | (1) 0.5%<br>(0.01, 2.9) | - |
|  | 6 µg with Algel-IMDG | - | - |  | - |
| **Total** |  | (33) 8.7%<br>(6.1, 11.9) | (7) 1.8%<br>(0.7, 3.8) | (34) 8.9%<br>(6.3, 12.3) | (2) 0.5%<br>(0.13, 3.8) |

*The groups received 3 µg with Algel-IMDG or 6 µg with Algel-IMDG. Data are shown as the number of participants who experienced an event (%) after receiving either dose 1 (0-7) or dose 2 (28-35 days). The grading scale for most adverse events was based on the FDA guidance document for the Toxicity Grading Scale for Healthy Adult and Adolescent Volunteers Enrolled in Preventive Vaccine Clinical Trials. For adverse events where grading was not described in the FDA guidance document, we used the Common Terminology Criteria for Adverse Events (CTCAE) grading.*

### Safety

A total of 6 (28·6%) out of 21 unsolicited adverse events were reported to be related to the vaccine. No significant difference was observed between the groups (Supplementary Table S7). The evaluation of severity grading and the relationship to the vaccine are described in Supplementary Table S8. No symptomatic SARS-CoV-2 infections were reported between days 0 and 75. However, the follow-up of routine SARS-CoV-2 nucleic acid testing was not conducted at any scheduled or illness visit. No serious adverse events were reported until day 56.

#### Phase 1 (at day 104, three months after the second dose)

No new solicited/unsolicited adverse events that occurred after day 42 were considered to be related to the vaccine by the investigators. No new serious adverse events were reported.

One case of symptomatic COVID-19 was reported in the Algel alone (control/placebo) group. The participant was screened on July 15^th^ and vaccinated on July 17^th^. The participant was unable to be contacted for the second vaccination visit and was considered to be lost to follow-up. The participant visited the site on November 27^th^ with complaints of chronic anosmia and a history of a positive SARS-CoV-2 rapid antigen test on August 16^th^.

## Discussion

We report interim findings from the phase 2 clinical trial of BBV152, a whole-virion inactivated SARS-CoV-2 vaccine. Both humoral and cell-mediated responses were observed. No neutralising antibody differences were observed between sexes and across age groups, albeit small numbers of participants were included in the ≥12-<18 and ≥55-<65 age groups. The vaccine was well tolerated in both dose groups with no serious adverse events.

The most common adverse event was pain at the injection site, followed by headache, fatigue, and fever. No severe or life threatening (Grade 4 and 5) solicited adverse events were reported. After any dose, the combined incidence rate of local and systemic adverse events in this study is noticeably lower than the rates for other SARS-CoV-2 vaccine platform candidates ^4,11-15^ and comparable to the rates for other inactivated SARS-CoV-2 vaccine candidates ^5,16^.

BBV152 induced binding (to both spike- and nucleocapsid protein epitopes) and neutralising antibody responses that were similar to those induced by other SARS-CoV-2 inactivated vaccine candidates ^5,16^. The current literature reports the variable persistence of humoral and cell-mediated responses acquired from natural infection ^17,18^. In the phase 1 trial, we evaluated an accelerated schedule (vaccination occurring two weeks apart). At day 104 (three months after the second vaccination dose), we observed detectable humoral and cell-mediated responses. Serum neutralising antibodies were detected in all the participants on day 104. These findings are in accordance with those on the mRNA-1273 vaccine, which will be licensed soon ^19^. A sizeable T cell memory population was also observed at this time point. A routine schedule (vaccination occurring four weeks apart) was evaluated in the phase 2 trial for 3 µg and 6 µg with Algel-IMDG. Here, immune responses were significantly higher than those in the phase 1 trial, which concurs with reports that a routine schedule offers higher immune responses ^20^. It is hypothesised that the humoral and cell-mediated responses reported in this study may persist until at least 6-12 months after the second vaccination dose.

An imidazoquinoline molecule (IMDG), which is a TLR7/8 agonist, has been used to augment cell-mediated responses ^21,22^. BBV152 is a whole-virion inactivated SARS-CoV-2 vaccine adjuvanted with Algel-IMDG. Both formulations were Th1-skewed with IgG1/IgG4 ratios above 1. The ratio of Th1/Th2 cytokines was clearly biased to a Th1 response with increased IFN-γ generation.

In the present study, BBV152 induced T cell memory responses, which was demonstrated by an increased frequency of antigen-specific CD4^+^ T cells expressing the memory phenotype marker CD45RO^+^. The increase in the CD4^+^CD45RO^+^CD27^+^ population also demonstrates the activation of the co-stimulatory marker CD27 and confirms the antigen recall memory T cell response. Further, the effector function of these cells was supported by the Th1-biased cytokine secretion observed on day 3. These results further corroborate our phase 1 results, where we reported an increased frequency of CD4^+^ T lymphocytes producing IFN-γ in Algel-IMDG recipients. The ability to secrete spike-specific IgG antibodies further demonstrates the long-lived memory response generated by BBV152. Similar findings supporting long-term immunity were reported by Sekine et al. in convalescent COVID-19 patients ^23^. Cell-mediated responses to other SARS-CoV-2 inactivated vaccine candidates have not been reported thus far.

This study was conducted in a time of rapid increases in daily diagnoses of COVID-19 cases. Among all participants who were screened, 48 (5.2%) and 63 (13.4%) reported positive SARS-CoV-2 nucleic acid tests and serology, respectively. In the phase 1 Algel alone (control arm) recipients, seroconversion was reported in 8·2% (1·9, 14·5), 18·1% (10·1, 29·3), and 32·9% (22·3, 44·9) on days 28, 42, and 104, respectively. At day 104, a total of 39 (52%) participants (receiving Algel alone) reported a 2-fold change in neutralising antibody titres. This suggests that both phase 1 and 2 trials are being conducted during a period of high ongoing SARS-CoV-2 circulation. In phase 2, no COVID-19 cases were reported from either group, while there was one cases of symptomatic COVID-19 in the control group of the phase 1 trial.

The results reported here do not permit efficacy assessments. The evaluation of safety outcomes requires extensive phase 3 clinical trials. We were unable to assess other immune responses (binding antibody and cell-mediated responses) of convalescent serum due to the limited quantity. No additional data on the severity of disease from symptomatic individuals were obtained. Last, this study population lacked ethnic diversity, further underscoring the importance of evaluating BBV152 in other populations. Longitudinal follow-up is important and is ongoing.

However, this study had several strengths. To ensure generalizability, this study was conducted with participants from diverse geographic locations, enrolling 380 participants across nine hospitals. The study enrolled participants with a wide range of ages and found no differences in immune responses across age groups. The overall participant retention rates were 96·8% and 93·2% in the 3 µg and 6 µg with Algel-IMDG groups, respectively.

Based on follow-up data from the phase 1 trial, at day 104 (three months after the second dose), despite a marginal expected decline in neutralising antibody titres, BBV152 has exhibited the potential to provide durable humoral immunity and cell-mediated immunity. From the phase 2 trial, the 6 µg with Algel-IMDG formulation was selected for the phase 3 efficacy trial, which is being carried out in 25,800 volunteers (NCT04641481).

## Data Availability

The study protocol is provided in the Supplemental Appendix Individual participant (de-identified) data will be made available when the trial is complete upon request directed to the corresponding author, after the approval of a proposal, data can be shared through a secure online platform.

## Acknowledgements

We would like to sincerely thank the principal and co-principal investigators, study coordinators and health care workers who were involved in this study. We express our gratitude to Dr. Sivasankar Baalasubramaniam from Indoor Biotechnologies, Bangalore, who assisted with cell-mediated response analyses. A special thanks to Drs. Arjun Dang and Leena Chatterjee of Dr. Dang’s Laboratory, which was the central laboratory for clinical laboratory testing. We appreciate the guidance from Dr. William Blackwelder on sample size estimation and statistical analysis planning. Drs. Shashi Kanth Muni, Sapan Kumar Behera, Jagadish Kumar, Vinay Aileni, and Ms. Akhila Naidu and Sandya Rani of Bharat Biotech participated in protocol design and clinical trial monitoring. Last, we thank the DSMB members (Drs. Kiran Kumar, Kiran Kishore, Srinivasa Rao, Sudha Madhuri, and Mr. Naradamuni Naidu) for their continued support and guidance on this ongoing clinical study. This vaccine candidate could not have been developed without the efforts of Bharat Biotech’s Manufacturing, Quality Control teams. All authors would like to express their gratitude to all frontline health care workers during this pandemic.

## Author Contributions

All listed authors met the criteria for authorship set forth by the International Committee for Medical Editors and have no conflicts to disclose. E.R. and K.M.V. accessed and verified the data (the CRO was responsible for generating the report). J.H., D.D., D.R., U.P., B.G., P.Y., and G.S. performed the immunogenicity experiments. K.M.V., P.S., S.R., V.S., and E.R. contributed to the analysis and manuscript preparation. S.R. was the study coordinator and helped immensely with the protocol design and interim report generation. P.A., S.P., A.P., N.G., and B.B. of NIV and ICMR, India, contributed various neutralising antibody assays and participated in writing this manuscript. All principal investigators were involved in the scientific review of this manuscript.

## References

1. WHO Coronavirus Disease (COVID-19) Dashboard. Available at https://covid19.who.int/.

2. World Health Organization DloC-cv, accessed on Oct 28th, 2020. Available at https://www.who.int/publications/m/item/draft-landscape-of-covid-19-candidate-vaccines.

3. Polack FP, Thomas SJ, Kitchin N, et al. Safety and Efficacy of the BNT162b2 mRNA Covid-19 Vaccine. New England Journal of Medicine 2020.

4. Jackson LA, Anderson EJ, Rouphael NG, et al. An mRNA Vaccine against SARS-CoV-2 — Preliminary Report. New England Journal of Medicine 2020.

5. Xia S, Duan K, Zhang Y, et al. Effect of an Inactivated Vaccine Against SARS-CoV-2 on Safety and Immunogenicity Outcomes: Interim Analysis of 2 Randomized Clinical Trials. JAMA 2020; 324(10): 951–60.

6. Logunov DY, Dolzhikova IV, Zubkova OV, et al. Safety and immunogenicity of an rAd26 and rAd5 vector-based heterologous prime-boost COVID-19 vaccine in two formulations: two open, non-randomised phase 1/2 studies from Russia. The Lancet 2020; 396(10255): 887–97.

7. Sarkale P, Patil S, Yadav P, et al. First isolation of SARS-CoV-2 from clinical samples in India. Indian Journal of Medical Research 2020; 151(2): 244–50.

8. Ganneru B, Jogdand H, Dharam VK, et al. Evaluation of Safety and Immunogenicity of an Adjuvanted, TH-1 Skewed, Whole Virion InactivatedSARS-CoV-2 Vaccine - BBV152. bioRxiv 2020: 2020.09.09.285445.

9. Pragya Yadav, Raches Ella, Sanjay Kumar et al. Remarkable immunogenicity and protective efficacy of BBV152, an inactivated SARS-CoV-2 vaccine in rhesus macaques, 10 September 2020, PREPRINT (Version 1) available at Research Square [+https://doi.org/10.21203/rs.3.rs-65715/v1+], Accesed on Nov 20, 2020.

10. Sreelekshmy Mohandas, Pragya D Yadav, Anita Shete et al. Immunogenicity and protective efficacy of BBV152: a whole virion inactivated SARS CoV-2 vaccine in the Syrian hamster model, 16 September 2020, PREPRINT (Version 1) available at Research Square [+https://doi.org/10.21203/rs.3.rs-76768/v1+]. Accessed on Nov 20, 2020.

11. Mulligan MJ, Lyke KE, Kitchin N, et al. Phase 1/2 study of COVID-19 RNA vaccine BNT162b1 in adults. Nature 2020.

12. Folegatti PM, Ewer KJ, Aley PK, et al. Safety and immunogenicity of the ChAdOx1 nCoV-19 vaccine against SARS-CoV-2: a preliminary report of a phase 1/2, single-blind, randomised controlled trial. The Lancet 2020; 396(10249): 467–78.

13. Zhu F-C, Li Y-H, Guan X-H, et al. Safety, tolerability, and immunogenicity of a recombinant adenovirus type-5 vectored COVID-19 vaccine: a dose-escalation, open-label, non-randomised, first-in-human trial. The Lancet 2020; 395(10240): 1845–54.

14. Zhu F-C, Guan X-H, Li Y-H, et al. Immunogenicity and safety of a recombinant adenovirus type-5-vectored COVID-19 vaccine in healthy adults aged 18 years or older: a randomised, double-blind, placebo-controlled, phase 2 trial. The Lancet 2020; 396(10249): 479–88.

15. Walsh EE, Frenck RW, Falsey AR, et al. Safety and Immunogenicity of Two RNA-Based Covid-19 Vaccine Candidates. New England Journal of Medicine 2020.

16. Zhang Y-J, Zeng G, Pan H-X, et al. Immunogenicity and Safety of a SARS-CoV-2 Inactivated Vaccine in Healthy Adults Aged 18-59 years: Report of the Randomized, Double-blind, and Placebo-controlled Phase 2 Clinical Trial. medRxiv 2020: 2020.07.31.20161216.

17. Gudbjartsson DF, Norddahl GL, Melsted P, et al. Humoral Immune Response to SARS-CoV-2 in Iceland. New England Journal of Medicine 2020; 383(18): 1724–34.

18. Dan JM, Mateus J, Kato Y, et al. Immunological memory to SARS-CoV-2 assessed for greater than six months after infection. bioRxiv 2020: 2020.11.15.383323.

19. Widge AT, Rouphael NG, Jackson LA, et al. Durability of Responses after SARS-CoV-2 mRNA-1273 Vaccination. New England Journal of Medicine 2020.

20. Xia S, Zhang Y, Wang Y, et al. Safety and immunogenicity of an inactivated SARS-CoV-2 vaccine, BBIBP-CorV: a randomised, double-blind, placebo-controlled, phase 1/2 trial. The Lancet Infectious Diseases.

21. Philbin VJ, Dowling DJ, Gallington LC, et al. Imidazoquinoline Toll-like receptor 8 agonists activate human newborn monocytes and dendritic cells through adenosine-refractory and caspase-1-dependent pathways. J Allergy Clin Immunol 2012; 130(1): 195-204.e9.

22. Shukla NM, Salunke DB, Balakrishna R, Mutz CA, Malladi SS, David SA. Potent Adjuvanticity of a Pure TLR7-Agonistic Imidazoquinoline Dendrimer. PLOS ONE 2012; 7(8): e43612.

23. Sekine T, Perez-Potti A, Rivera-Ballesteros O, et al. Robust T Cell Immunity in Convalescent Individuals with Asymptomatic or Mild COVID-19. Cell 2020; 183(1): 158-68.e14.

